# Adaptations and interpretation of Intelligence Tests for Adults with Visual Impairment: A Literature Review and Interviews with Healthcare Professionals

**DOI:** 10.64898/2026.03.11.26347170

**Authors:** Cynthia Lamper, Marjolein L.A. Onnink, Marit van Buijsen, Hilde P.A. van der Aa, Edine P.J. van Munster

## Abstract

**Background:** Administering and interpreting intelligence tests for adults with visual impairment (VI) presents practical and methodological challenges, because standard test batteries rely heavily on visual tasks and lack specific norm-groups tailored to this population. This study examined practical adaptations and interpretation strategies currently used in intelligence testing based on international literature and input from Dutch low vision service (LVS) providers.

**Methods:** A mixed-method design was applied. An exploratory literature review was conducted using PubMed, PsycINFO, Google Scholar and Web of Science to identify studies published in the past ten years addressing adaptations and/or interpretation of intelligence tests. Additionally, semi-structured interviews were held with nine healthcare professionals (HCPs) from Dutch LVS organizations to explore current practices and perceived needs.

**Results:** Eight publications met inclusion criteria. All reported use of intelligence tests; six described task adaptations, which varied considerably. One addressed interpretation of results for adults with VI. None reported norm-groups adapted for people with VI.

Interviews highlighted challenges due to absence of accessible tests, leading HCPs to solely rely on the assessment of verbal subtests, mainly of the Wechsler Adult Intelligence Scale – Fourth Edition (WAIS-IV) or making adaptations to the assessment of performance tasks for observational purposes. Approaches to administering tests varied widely.

**Conclusion:** Both literature and interviews with HCPs indicate that no specific intelligence tests tailored for adults with VI have been developed in the past decade, making accurate Total Intelligence Quotient (TIQ) measurement currently impossible. There is an urgent need for tailored tests with corresponding norm-groups and/or standardized protocols to ensure validity and consistency. Until then, HCPs should select predominantly verbal subtests tailored to the individual, observe performance closely, and explicitly report considerations.

**Strengths and limitations of this study:**

- Mixed-methods design combining an exploratory literature review with qualitative interviews enabled methodological triangulation and a comprehensive understanding of current practices.
- Systematic and transparent review procedures, including dual independent screening, use of JBI critical appraisal tools, and PROSPERO registration, enhanced methodological rigor.
- A final strength was the use of established qualitative data collection and analysis approaches (semi-structured interviews, reflexive thematic analysis, and double coding), which enhanced the depth and credibility of the findings.
- The exploratory (non-systematic) nature of the literature review, including broad database searches and absence of a fully reproducible search strategy, may limit reproducibility and increase the risk of selection bias.
- Lastly, qualitative sampling through snowball recruitment within a single national context (Dutch LVS) and a relatively small sample size may limit transferability and increase the risk of selection bias.

**Key messages:**

- Recent literature on intelligence testing in adults with Visual Impairment (VI) provides no evidence of valid and reliable tools for accurately measuring Total Intelligence Quotient (TIQ);
- Healthcare professionals (HCPs) working with adults with VI primarily administer verbal subtests from the Wechsler Adult Intelligence Scale - Fourth Edition (WAIS-IV);
- More research should be performed on the development of specific intelligence tests and adapted norm-groups tailored to adults with VI to assess valid TIQ.

## Introduction

In the general population, around 3 to 4% of adults report difficulties with vision or activities requiring sight (1). According to the International Classification of Diseases, vision loss is classified as visual impairment (VI) whenever a person has a visual acuity of 6/12 or lower, meaning that this person can read at 6 meters what people with normal vision can read at twelve meters. VI is categorized into mild VI (6/12 to 6/18), moderate VI (6/18 to 6/60), severe VI (6/60 to 3/60), and blindness (3/60 or lower) (2). Moreover, VI may be congenital or acquired later in life. Prevalence rates are considerably higher in specific subgroups, for example in adults with intellectual disabilities 15% has VI and even 5% is blind (1, 3). In the Netherlands, more than 300,000 people are living with a VI and this number is expected to increase in the coming years (4, 5).

People with VI experience more difficulties in performing (instrumental) activities of daily living (6). Examples include mobility-related activities, driving, reading, performing computer work, and engaging in leisure activities. These limitations can result in reduced autonomy in daily life and can restrict participation in work-related and social activities (6–10). When an adult with VI has a need for support or requires assistance with activities of daily living, it is often essential to gain understanding of their cognitive functioning (11). This understanding can provide insight into the person’s capacity to learn new skills and the most effective ways to support their learning and participation in work and social activities (12–14).

According to the Cattell–Horn–Carroll (CHC) theory, cognitive abilities are structured hierarchically into a general intelligence factor, consisting of several broad abilities such as fluid reasoning, crystallized knowledge, visual–spatial processing, working memory, and processing speed, and numerous more specific abilities (15). Intelligence tests play a fundamental role in neuropsychological practice, as they provide a structured assessment of these cognitive abilities (16). These tests provides insight into various indices of intelligence, leading to a distinction between strong and weak cognitive functions (17, 18). Identifying strengths and limitations through testing helps tailor interventions, support strategies, and decision-making in healthcare. Specific information from intelligence testing is essential for diagnosis, supporting indications, formulating treatment plans, and enhancing interdisciplinary communication (19). Therefore, they are essential for LVS providers. However, to date, no specific guidelines appear to be available.

Dutch HCPs within low vision service organizations seem to express a need for clear protocols to improve the accuracy in the assessment of intelligence tests. To our knowledge, the last literature overview of intelligence tests was published in 2019 by Rączy & Korczyk (20). While these examples illustrate efforts to adapt the assessment of intelligence tests for this population, they primarily target children or remain limited in scope and psychometric quality. Consequently, there is currently no standardized, validated intelligence test available for adults with VI, highlighting a critical gap in assessment tools for this group. Therefore, this study aims to get insight into the currently used adaptations made during the assessment of intelligence tests and interpretation of the results of these tests for adults with VI in international literature and in Dutch LVS.

## Methods

In this mixed-methods study, a literature review and an interview study among HCPs of LVS were conducted. The literature review was registered at PROSPERO number: CRD42024613453 (PROSPERO). For the interview study, verbal informed consent was obtained from all HCPs before the start of the interview. All HCPs provided recorded informed consent for participation in the study and for the use of their data for research purposes. This manuscript was developed in accordance with the Standards for QUality Improvement Reporting Excellence (SQUIRE 2.0) guidelines to ensure transparent and comprehensive reporting (21).

### Literature review

#### Search strategy and in- and exclusion criteria

An exploratory literature review was conducted to investigate existing research, focusing on adults with VI who mentioned adaptations made in intelligence testing and/or the interpretation of the results of intelligence tests in adults with VI. Searches were performed in the databases PubMed, PsycINFO, Google Scholar and Web of Science to identify publications between November 1^st^ 2014 and November 1^st^ (Supplementary File 1). Publications were eligible for inclusion if they addressed adaptation(s) or interpretation of intelligence tests in adults with VI. Intelligence tests were defined as assessments designed to calculate a total Intelligence Quotient (IQ). Publications in all languages were considered; when necessary, Artificial Intelligence tools (ChatGPT and Copilot) were used for translation. Because intelligence tests are designed to assess broad cognitive functioning, publications focusing exclusively on cognitive tests were excluded from the review (15). Moreover, studies with populations consisting solely of children (under age 18) or adults with dual sensory loss (hearing and vision impairment) were excluded.

#### Selection process

The literature search generated a total of 3,006 publications: 2,521 in PubMed, 298 in PsycINFO, 65 in Google Scholar, and 39 in Web of Science. After removing duplicates, three reviewers used Rayyan to independently screen 2,984 publications on title and abstracts for eligibility (C.L.: all titles and abstracts, E.M. and M.B.: both half of the titles and abstracts). Thereafter, 29 Full Text Publications (FTP) were checked by two reviewers (C.L.: all FTP, E.M., and M.B.: both half of the FTP). If conflicts arose about the in- or exclusion of a publication, these were discussed, and if no consensus was reached, the third and fourth reviewer (M.B. and M.O.) were involved and performed an independent screening. After screening, eight publications were included in this review.

**Figure 1.**
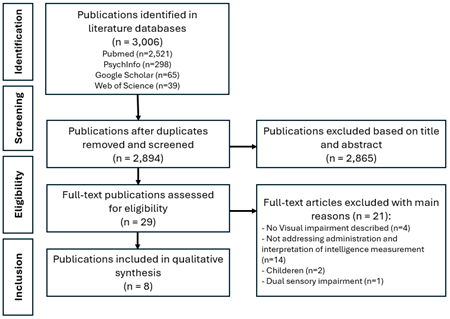
Flow diagram of selection process.

#### Quality assessment

To assess the quality of the included publications, critical appraisal tools published by Joanna Briggs Institute (JBI) were applied (22). These tools evaluate key aspects related to the reliability, relevance, and results of the publications. The most suitable evaluation tool was selected for each type of publication, *e.g.* reviews (23), case control studies (24), cross sectional studies (24), case studies (25), cohort studies (24), and case reports (24). Each study was independently assessed by two reviewers (C.L. and M.O.). Discrepancies were discussed, and if necessary, a third reviewer (E.M.) was consulted. The results of the quality assessment informed the interpretation of the overall strength of the evidence.

When in the JBI tools was asked about ‘exposure measured,’ it was interpreted as the tests to which people were exposed. According to a systematic review of practice use of JBI critical appraisals, to indicate an overall quality ‘score’, the total number of ‘yes’ responses was counted and this was transferred to a percentage (26). A quality assessment was then performed, with the classifications low (<50%), adequate (50–75%), or good (>75%).

#### Analysis

The following characteristics were extracted for each publication: author, year of publication, study design, used intelligence test, adaptations made for people with VI, interpretation of test scores, and additional main findings relevant to the research question. In addition, psychometric properties of the instruments that were used, such as reliability and validity specifically for individuals with VI, were reported. The extracted data were organized in Microsoft Excel (version 25) to facilitate the analysis, to enable a structured comparison across publications, and to identify patterns in the use and adaptation of intelligence tests for adults with VI.

### Interviews with HCPs

#### Sample and recruitment

To gain -in-depth insights into intelligence testing among adults with VI, -semi structured qualitative interviews were conducted with HCPs working at one of three Dutch LVS locations between June 16, 2025, and October 6, 2025.HCPs were eligible for this study when they were aged 18 years or older, work at one of the three Dutch LVS, were involved in psychodiagnostics testing of adults with VI, and spoke and wrote Dutch fluently. Recruitment was carried out using a snowball sampling approach, whereby HCPs were referred through the National expert network on VI and Psychiatry in the Consortium for Expertise in living with VI.

#### Data collection

All interviews took place within the secure environments of Microsoft Teams and Apple Voice Memos, and were audio recorded. Audio files were transcribed verbatim by Amberscript (27). Personal and identifiable data were removed from transcripts to maintain confidentiality. The interviews followed a semi-structured topic guide, which was developed based on existing literature, input from LVS behavioral scientists, the National expert network on VI and Psychiatry and two experts by experience. The topic guide included themes such as the suitability of currently used tests, challenges and needs in administering intelligence tests, standardization within teams, interpretation of test results, and recommendations for adaptations and implementation in practice.

After the topic was introduced, the interview proceeded with several theme-related questions, for example: the background and experience of the HCP; the use of intelligence tests in adults with VI; adaptations and methodologies for assessing the intelligence of individuals with VI; approaches to interpretation and norming of these tests; and, finally, the need for standardized protocols and the further development of intelligence tests specifically designed for adults with VI. Figure 2 presents example topic guide questions. The complete interview guide is provided in Supplementary File 2. Interviews were conducted by a female PhD-candidate/ Licensed Health Care Psychologist with expertise in intelligence testing (M.O.) Data collection continued until no new concepts were discussed by interviewees (28).

**Figure 2.** Examples of topic guide questions. - What is your experience with the use of intelligence assessments in individuals with VI?
- And what differences do you observe compared to sighted individuals?
- Which intelligence assessments are utilized? How frequently?
- Which adaptations do you make, or would you make, in the administration of intelligence assessments for individuals with VI?
- What are your experiences with tactile or verbal alternatives (such as Braille versions or verbal subtests) in intelligence assessment?

#### Data analysis

The transcripts of all interviews were analysed using reflexive, inductive, thematic analysis to identify and describe the main themes that were indicated in the interviews by HCPs in the use, adaptation, and interpretation of intelligence tests (28). All analyses were performed by two researchers (C.L. and M.O.) by using MAXQDA software (29). Three transcripts (33%) were independently double-coded by two researchers (C.L. and M.O.). They applied codes to the text with line-by-line coding, then discussed emerging codes and themes and generated the coding framework. M.O. then applied the coding framework to all remaining transcripts (n=6), adding new codes as transcripts became available. This process was followed until no new codes were identified and data saturation was achieved. These new codes were discussed with C.L. As a cross-check, C.L. conducted additional analyses on parts of three interviews (33%) selected through random sampling. Codes were then arranged into themes pertaining to the research questions by M.O. and C.L. The research team (C.L., E.M., M.B., M.O. and H.A.) finalized themes and approved the final report.

## Results

### Literature review

#### Study characteristics

Eight publications were included in this review (Table 1). The publications consisted of one review (30), two case control studies (31, 32), two cross-sectional studies (33, 34), two cohort studies (35, 36) and one case-report (37). The review study of Galiano et al. [2018] was translated from French to Dutch using Artificial Intelligence (ChatGPT) (30). All publications reported the intelligence test used (30–37), six publications mentioned how the intelligence test was adapted for people with VI (30, 32–34, 36, 37), and only one publication reported on the interpretation of the test scores (34).

**Table 1.** Characteristics of included publications.

| Author (Year) | Study Design | Study sample (N) | Intelligence Test | Adaptation | Interpretation |
| --- | --- | --- | --- | --- | --- |
| Bischoff et al. [2015] (35) | A longitudinal natural history study | N=19 WFS-participants<br>N=25 T1DM-participant<br>N=25 No VI | WASI, Letter-Number Sequencing (part of WAIS-IV, Digit Span (part of WAIS-IV). These subtests were used to generate a VIQ. | n.d.a. | n.d.a. |
| Dang Do et al. [2022] (36) | Cohort study<br>Φ | N=22 participants<br>CLN3 | WAIS-IV | VIQ was reported as nonverbal testing was not feasible | n.d.a. |
| Galiano et al. [2018] | Review | - | WAIS-IV, B-101 DV, HISB, Stanford-Ohwaki-Kohs Tactile Block Design | For the WAIS-IV psychologists used verbal subtests or | n.d.a |
| (30) |  |  | Intelligence Test for the Blind, TONI | subtests focused on working memory index. |  |
| de Haan et al. [2020] (31) | Simulation study with case-control design $\Phi$ | N=238<br>Consisting of:<br>N=117 simulated cases for VI<br>N=121 controls | DART | n.d.a. | n.d.a. |
| Merolla et al. [2022] (37) | Case report $\Phi$ | N=1 with VI caused by Boucher-Neuhäuser Syndrome | Digit Span test, test from the Working Memory Scale of the WAIS-IV, and tests from the Verbal Comprehension Scale of the WAIS. | A neuropsychological battery of tests that required ad hoc selection was compounded, and in some standardized tests were adjusted in order to bypass visual and motor coordination deficits. | n.d.a. |
| Nitzan et al. [2024] (33) | Retrospective cross-sectional study | N=1,410,616 | General Intelligence Score consisting of: Otis Classification Test-Revised, Similarities-R, Arithmetic-R, and Raven's Progressive Matrices-R | Examinees were instructed to bring their prescribed optical correction devices, if applicable, for the duration of the assessment. | n.d.a. |
| Pigeon et al. [2015] (34) | Cross-sectional design $\Phi$ | N=48<br>Consisting of:<br>N=14 early blind<br>N=10 late blind<br>N=24 No VI, who were blindfolded during testing | Six subtests from the verbal portion of the WAIS-IV;<br>1) Digit Span test; 2) N-back test; 3) Plus-minus task; 4) Selective attention test; 5) Sustained attention test; 6) divided attention test | Different attentional components of the participants were assessed with tests that were standardized or designed for the study because no test was available for persons with VI. An auditory version of the plus-minus task was used to assess attentional switching. | It is important to note the presence of a ceiling effect only for the blind participants in the present study (this effect was observed in 6 early and 2 late blind participants in the forward span test and 3 early blind participants in the backward span). |
| Wright et al. [2024] (32) | Prospective study with case-control design $\Phi$ | N=63<br>Consisting of: N=21 with CRB1 retinopathy , N=42 with vision loss due to other inherited | Digit Span test (part of WAIS-IV) | To account for VI, cognitive function was assessed using standardized neuropsychological tests that did not involve visual processing and required only verbal | n.d.a. |
|  |  | ocular<br>conditions |  | interaction. |  |
| N = number<br>WFS = Wolfram Syndrome<br>T1DM = Type one Diabetes Mellitus<br>VI = Visual Impairment<br>WASI = Wechsler Abbreviated Scale of Intelligence<br>WAIS-IV = Wechsler Adult Intelligence Scale - Fourth Edition<br>VIQ = Verbal Intelligence Quotient<br>n.d.a. = No Data Available<br>CLN3 = Ceroid Lipofuscinosis, Neuronal, type 3<br>B-101 DV = Bonnardel 101 – Déficients Visuels<br>HISB = Haptic Intelligence Scale for the Blind<br>TONI = Tactile Test of Nonverbal Intelligence<br>DART = Dutch Adult Reading Test<br>CRB1 = gene Crumbs homolog 1 retinopathy<br>Φ = not described in the article but interpreted by the authors. |  |  |  |  |  |

#### Used intelligence tests

The Wechsler Adult Intelligence Scale-IV (WAIS-IV) was the most frequently applied instrument, reported in four publications. Some publications only used subtests of the WAIS-IV, such as, Digit Span (N=4) and Letter–Number Sequencing (N=1). Other instruments comprised the Wechsler Abbreviated Scale of Intelligence (35), the Dutch Adult Reading Test (31) and specialized tactile measures such as the Stanford–Ohwaki–Kohs Tactile Block Design Intelligence Test and the TONI–Tactile Test of Nonverbal Intelligence (30). Additionally, Nitzan et al. (2024) used a composite General Intelligence Score based on Otis Classification Test-Revised (OTIS-R), Similarities-R, Arithmetic-R, and Raven’s Progressive Matrices-R (33). For an overview of the mentioned IQ tests, see Supplementary File 3.

#### Adaptations for adults with VI

Adaptations that were made in the assessment varied considerably across publications. Two publications reported administering only the verbal subtests of the WAIS-IV due to the infeasibility of visual components (30, 36). Other adjustments included the use of enlarged materials, modified lighting conditions, and auditory versions of tasks, (33, 34, 36, 37). An example of auditory tasks is the plus–minus task, which measures working memory and mental flexibility by having participants add and subtract numbers in their mind. Merolla et al. (2022) described ad hoc modifications to bypass visual and motor coordination deficits (37); *i.e.* by presenting cognitive tasks through the auditory verbal modality and selecting a balanced set of subtests that minimized visual and motor demands. The review by Galiano et al. (2018) highlighted the existence of haptic alternatives, including the Haptic Intelligence Scale for the Blind (HISB) and tactile block design tests, although these instruments appear to be rarely implemented in current practice (30).

#### Interpretation of test scores

Only one publication addressed the interpretation of intelligence test scores for adults with visual impairments (34). Authors reported a ceiling effect among blind participants on forward and backward span tests and suggested superior working memory based on N-back results (measures working memory and attention by testing how well participants can update and recall information), though comparability with other studies was limited. The remaining seven publications provided no guidance on interpretation, and none reported using adapted normative data.

#### Quality assessment

A detailed explanation of the quality assessment can be found in Supplementary file 4. Quality scores ranged from 27% to 100%, with corresponding conclusions categorized as Low (37,5%, Adequate (37,5%), or Good (25%).

Overall, the methodological quality of the included studies varied considerably. Reviews showed the lowest quality. Cohort studies demonstrated mixed quality, ranging from adequate to low. Case-control studies displayed substantial variation, with one high-quality and one low-quality study. The single case report was of adequate quality. Cross-sectional studies also varied, with quality assessments ranging from good to adequate.

Overall, case-control and cross-sectional designs tended to achieve higher quality scores compared to reviews and cohort studies. Table 2 summarizes the individual study characteristics and quality ratings.

**Table 2:** Quality assessment of each included study using the JBI Critical Appraisal Checklists per study design.

| Author (year) | Study design | Score (%) | Conclusion |
| --- | --- | --- | --- |
| Bischoff et al. [2015] (35) | Cohort study | 7/11 (64%) | Adequate |
| Dang Do et al. [2022] (36) | Cohort study | 4/11 (36%) | Low |
| Galiano et al. [2018] (30) | Review | 3/11 (27%) | Low |
| de Haan et al. [2020] (31) | Case control study | 9/10 (90%) | Good |
| Merolla et al. [2022] (37) | Case report | 4/8 (50%) | Adequate |
| Nitzan et al. [2024] (33) | Cross-sectional study | 8/8 (100) | Good |
| Pigeon et al. [2015] (34) | Cross-sectional study | 5/8 (63%) | Adequate |
| Wright et al. [2024] (32) | Case control study | 4/10 (40%) | Low |

### Interviews with HCPs

A total of nine HCPs from LVS, *i.e.* Center A (N=3), Center B (N=3), and Center C (N=3) participated in the interview study. Of these HCPs, three held a background in pedagogical sciences, while six were trained in (developmental) psychology. Four HCPs had also completed postgraduate training as Licensed Health Care Psychologist (in Dutch: GZ-psychologist). On average, HCPs reported 16 years of experience administering intelligence tests (ranging from 5 to 31 years). The HCPs had on average 13 years (range: 1 to 31 years) experience with administering intelligence tests on adults with VI. Details of each HCP can be found in Table 3.

**Table 3:**
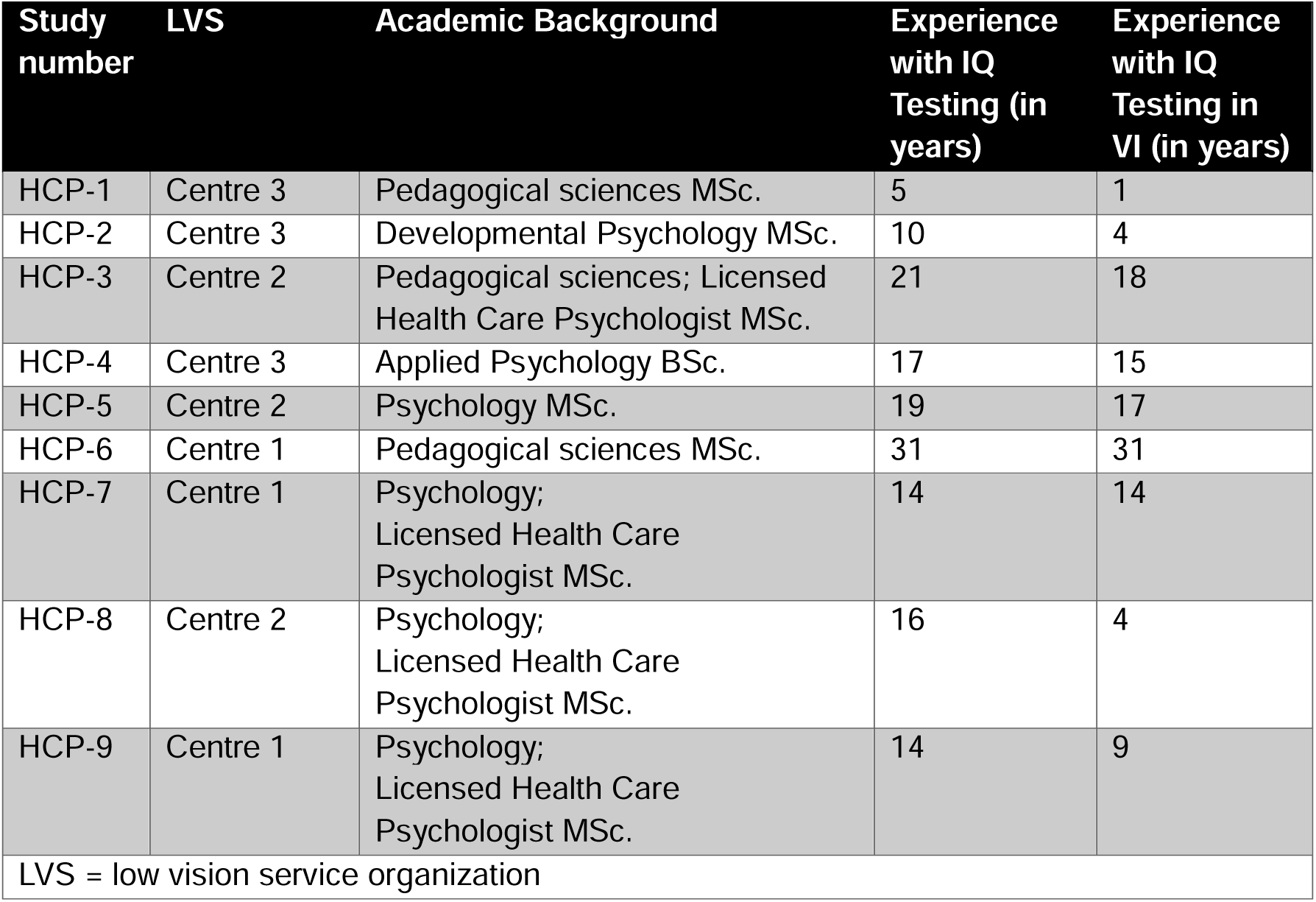
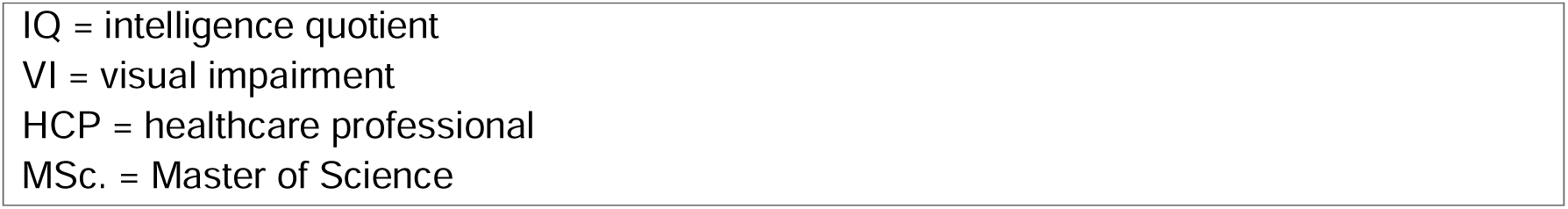
Background characteristics of included healthcare professionals.

| Study number | LVS | Academic Background | Experience with IQ Testing (in years) | Experience with IQ Testing in VI (in years) |
| --- | --- | --- | --- | --- |
| HCP-1 | Centre 3 | Pedagogical sciences MSc. | 5 | 1 |
| HCP-2 | Centre 3 | Developmental Psychology MSc. | 10 | 4 |
| HCP-3 | Centre 2 | Pedagogical sciences; Licensed Health Care Psychologist MSc. | 21 | 18 |
| HCP-4 | Centre 3 | Applied Psychology BSc. | 17 | 15 |
| HCP-5 | Centre 2 | Psychology MSc. | 19 | 17 |
| HCP-6 | Centre 1 | Pedagogical sciences MSc. | 31 | 31 |
| HCP-7 | Centre 1 | Psychology; Licensed Health Care Psychologist MSc. | 14 | 14 |
| HCP-8 | Centre 2 | Psychology; Licensed Health Care Psychologist MSc. | 16 | 4 |
| HCP-9 | Centre 1 | Psychology; Licensed Health Care Psychologist MSc. | 14 | 9 |
| LVS = low vision service organization |  |  |  |  |

IQ = intelligence quotient VI = visual impairment HCP = healthcare professional MSc. = Master of Science

The mean duration of the interviews was 24,22 min (range: 24,27 min to 44,49 min). The themes and subthemes identified through data analysis are presented in Table 4 and will be explained in more detail in the following paragraphs.

**Table 4:** Overview of themes and subthemes emerged during thematic analyses.

|  | Theme | Subtheme |
| --- | --- | --- |
| 1 | Differences in the administration of intelligence tests for adults with and without VI | • Background information<br>• Difference between VI and without VI<br>• Adaptations for VI |
| 2 | Consideration in test selections | • Used test(s)<br>• Alternative test to measure intelligence or cognition<br>• Difference between subgroups of adults with VI |
| 3 | Test administration | • Protocols and guidelines for measurement of TIQ of adults with VI<br>• Norm-groups |
| 4 | Analyses, report and feedback | • Interpretation of test results<br>• Validity, reliability and credibility of test results<br>• Feedback of test results to adults with VI<br>• Communication about test results |
| 5 | Needs and wishes | • Knowledge gaps<br>• Wishes for future developments in intelligence testing |
| VI= visual impairment<br>TIQ = total intelligence quotient |  |  |

#### Theme 1: Differences in the Administration of Intelligence Tests for Adults With and Without Visual Impairment

All HCPs reported notable differences in the administration of intelligence tests between adults with VI and those without. They frequently adapted testing procedures to accommodate the needs of adults with VI. A commonly cited issue was that vision loss is often too severe to conduct the non-verbal components of the intelligence test, resulting in only the verbal section being conducted. In some cases, HCPs attempted to explain visual items to clients with residual vision, or they employed magnification tools or adjusted lighting conditions. However, they observed that visual components of tests are often too fatiguing or demanding for clients.

Seven HCPs indicated that adults with VI needed more time to complete the tests compared to adults without VI. Furthermore, limited familiarity with academic skills and language-related aspects, such as idioms and proverbs, may have hindered their ability to provide appropriate responses. Non-verbal communication during testing was also conducted different from sighted people; HCPs noted that clients cannot be encouraged through non-verbal cues. One HCP mentioned using more verbal communication to make her presence known during the test administration.

##### HCP 2

> *“I think you make your presence known by using more language. When administering a test to individuals who are completely blind, you tend to verbalize more where you are, how you respond. Even though, according to standardized protocols, you are not supposed to do that.”*

One HCP emphasized the importance of minimizing adaptations during test administration to preserve the reliability and validity of the results as much as possible.

#### Theme 2: Considerations in Test Selection

##### Administered Tests and Alternatives

All HCPs reported experience with administering the WAIS to adults with VI. In cases where intellectual disability was suspected, the WISC or WPPSI were also used. Additionally, four HCPs mentioned familiarity with or even used the “Intelligentietest voor Visueel Gehandicapte Kinderen” (ITVIK) (Translated to English: Intelligence test for visually impaired children), which is a Dutch intelligence test developed for children with VI consisting of tactile components. However, HCPs mentioned this test is rarely used with adults, as its available norm groups were developed for children and therefore do not provide appropriate norms for adult populations. One participant also noted that the Vithoba Paknikar Performance Tests for the Blind, although specifically designed for this population, are no longer available because they are very outdated and no longer accessible (38). Five HCPs explicitly stated that they are not aware of any intelligence tests available in Braille.

Seven HCPs reported using components from other (sub)tests, such as the ADAPT, and Tactile Profile, to supplement the chosen intelligence test and obtain a broader understanding of the individual’s cognitive functioning.

###### HCP 5

> *“For example, if there is a suspected intellectual disability, I also administer the ADAPT to incorporate observed adaptive skills and provide a more comprehensive idea of an individual’s daily functioning.”*

##### Criteria for Test Selection

HCPs were asked about the criteria they use when selecting an intelligence test. They indicated that the availability of tests within the organization is a key factor. Other considerations included the referral question, whether the test is needed for diagnostic purposes, prior to testing history, and the client’s remaining vision. Two HCPs stated that they also consider the expected intellectual level of the client to avoid overburdening them and prevent negative experiences. Furthermore, the importance of clearly defining the purpose of the assessment before administering an intelligence test and weighing whether calculating a full-scale IQ is truly necessary were emphasized by two HCPs. Four HCPs also considered the client’s functional abilities in daily life and aimed to assess broader cognitive functioning.

##### Differentiation Within the Client Group

HCPs were asked whether they differentiated between the severity and tailored visual characteristics of the impairment when selecting a test. They reported to consider factors such as visual field size, acuity, percentage of vision, and daily functioning to determine whether subtests that rely on visual information could be administered and whether enlarged materials or additional time were needed. Furthermore, they took into account whether the VI is congenital or acquired, as this influenced how some of the visual items of a test were described. In example, individuals with prior visual experience may still perform tasks relying on visual memory, such as drawing a house, whereas such tasks are not applicable to those born blind, who lack visual referential frameworks. For adults with congenital blindness, descriptions of objects sometimes required adjustments due to differences in mental imagery, as they have no visual reference for concepts such as a picture of a house on fire.

###### HCP 1

> *“If someone has never seen, you have to describe things differently, because they don’t have a mental image of a tree or an elephant.”*

#### Theme 3: Test Administration

##### Procedures

Eight HCPs reported that they aim to create optimal conditions during test administration. This included selecting the most suitable time of day based on the client’s energy levels, optimizing the test room (e.g., light settings), dividing the test into multiple sessions if needed, clarifying questions, using enlarged materials, and allowing extra time for responses.

##### Protocols, Guidelines, and Norms

Three HCPs believed that no specific protocols or guidelines exist within their organization for administering intelligence tests to adults with VI. The remaining six were unsure. While the WAIS-IV is often used by default, this is not formally documented. Some HCPs applied existing guidelines developed for other (sighted) adult populations, such as adults with intellectual disabilities. HCPs emphasized that when additional time is given, it is important for the test administrator to be aware of its implications for score interpretation. Some HCPs followed the manual and recorded the standard time and score, but still allowed the client to complete the test (“testing to the limits”) to avoid frustration and provide a positive experience. They also stressed the importance of observing how the client approached the questions and estimated their processing speed. All HCPs expressed that having a protocol or guideline would be helpful to better document deviations during test administration.

###### HCP 9

> *“You encounter problems with visually-based tasks. That is the main issue. You constantly have to ask yourself: am I measuring what I intend to measure, or is the visual impairment dominating? If someone is blind, that is clear. But with low vision, it becomes a puzzle.”*

Eight HCPs specifically noted that no tailored norm-group exists for scoring intelligence in adults with VI. Some used the standard normative data provided in the test manuals, whereas other HCPs preferred to only use their observations. There was also skepticism by some about the feasibility of developing a norm-group for this population due to its heterogeneity.

###### HCP 2

> *“I am doubtful, because if you want a more precise IQ score, then yes, it [norm-groups] is necessary. But getting that done is difficult. A norm-group for blind and low-vision individuals might not capture everything. You might miss aspects of how the material is presented and how the person interacts with it. That gives you a lot of information.”*

##### Advantages and Disadvantages of Test Adaptations for VI

HCPs reported that they regularly implement adaptations when administering intelligence tests to adults with VI. A frequently mentioned reason is the ability to provide personalized assessment that reflects the client’s actual capabilities.

Adaptation often involved evaluating which components of the available intelligence tests are suitable for the client. In many cases, only the verbal subtests were administered.

###### HCP 7

> *“I mainly see the advantages of these adaptations. I am not sure what the limitations are. Are there any? Ultimately, you want the conditions to be as optimal as possible to conduct the test. If additional lighting helps, or anything else, I only see benefits in applying such measures.”*

HCPs mentioned that limiting the assessment to verbal components alone may not provide a thorough understanding of a client’s cognitive abilities. Intelligence is a multifaceted construct, and focusing solely on verbal reasoning may overlook other important domains such as processing speed, working memory, and non-verbal problem-solving.

Other important drawbacks highlighted by several HCPs were the reduced reliability and validity of the test results due to the adaptations. Moreover, the lack of standardized norms for adapted procedures was frequently mentioned as a limitation. Furthermore, HCPs noted that administering non-verbal subtests to clients with residual vision can be overly demanding and fatiguing, which may negatively affect concentration and performance.

#### Theme 4: Analysis, reporting and feedback

##### Interpretation

Eight HCPs indicated that a Total IQ (TIQ) score is typically not calculable when only verbal tasks are administered. This limitation may lead to under- or overestimation of a client’s abilities, particularly in cases of disharmonic profiles. For example, in clients who are verbally strong or have difficulty expressing themselves, insight into processing speed or other non-verbal domains may be difficult.

Five HCPs stressed the need for cautious and descriptive interpretation. All HCPs often included observations during test administration in the report to contextualize the results and highlight the impact of the adaptations. Three HCPs reported using standard norms for interpreting verbal components, but emphasized the importance of describing the likely influence of the VI, especially in the absence of tailored norms for this population.

###### HCP 2

> *“Interpretation requires caution and additional descriptive detail.”*

###### HCP 3

> *“You work with the information available. In certain groups, verbal scores may lead to overestimation, especially in individuals who are verbally overdeveloped; this is particularly true for those who are blind from birth. These factors must be carefully considered in the conclusions, although the information can still be valuable.”*

##### Reliability and Validity

All HCPs expressed concerns about the reduced reliability and validity of test results in adults with VI, particularly when visual tasks are tailored (e.g., enlarged stimuli or extended time, depending on an individual’s visual abilities). Three HCPs believed that verbal components are likely to remain reliable and valid, when provided standard testing conditions are met. However, one HCP noted that even verbal tasks may pose challenges, as clients may not recognize certain situations due to lack of visual experience, potentially affecting performance. Tasks such as digit span are considered less affected by visual limitations.

###### HCP 5

> *“The better the vision, the more reliable the results; the poorer the vision, the less reliable.”*

##### Reporting

All HCPs emphasized the importance of clearly and carefully documenting environmental conditions and all adaptations that are made during test administration. They suggested that observations regarding task approach and compensatory strategies (e.g. reduced viewing distance) should be included. Furthermore, the role of the VI and its impact on performance must be thoroughly described, along with any modifications such as lighting, presence of others, or extended time. One HCP recommended video recording to test administration and observations to ensure accurate documentation of procedures and interventions.

HCPs also noted that results should be interpreted with caution, and that definitive scores may not be appropriate. One HCP suggested limiting the validity period of the report, given that testing reflects a single moment in time.

###### HCP 3

> *“I am very used to interpreting and evaluating results with full awareness that the procedure deviates from the manual. You describe this and include it in the conclusions. These are standard phrases regarding reliability. You always mention that the measurement is not reliable due to the VI and explain how this has influenced the outcomes*.”

##### Feedback to Clients

Seven out of nine HCPs stated that feedback should be provided verbally. Sufficient time should be allocated to clarify how conclusions were made and to explain the role of the VI and adaptations during testing. In some cases, HCPs create a separate client report tailored to the client’s estimated cognitive level. As in standard practice, legal representatives also receive a formal report.

Several HCPs emphasized that feedback should be personalized and may benefit from including examples of test items to enhance client understanding. Ensuring that clients recognize themselves in the results is a key concern. Additionally, HCPs aim to leave clients with a positive impression by highlighting strengths.

###### HCP 7

> “*You remain transparent, but within that framework, you provide explanations and try to align with the client wherever possible.”*

#### Theme 5: Needs and Wishes

HCPs expressed a range of needs and wishes concerning the assessment of intelligence in adults with VI. All HCPs indicated a desire for norm-groups tailored to adults with VI, or alternatively, intelligence tests adapted to different levels of visual functioning. Several HCPs suggested that a standardized protocol could enhance reliability and reduce the influence of individual interpretation by ensuring consistency in test administration.

When discussing the wishes for future assessment of intelligence, several HCPs called for a dedicated manual that outlines procedures for describing functioning, handling test materials, and accounting for the impact of VI on test performance. One HCP also expressed the need for specialized training in administering tests to this population. However, multiple HCPs acknowledged the challenge of developing a new test due to the heterogeneity of VI and the differences between adults with congenital versus acquired VI.

##### HCP 3

> *“It would be ideal if we could work uniformly in both administration and interpretation. I am not sure norm-groups are feasible, but perhaps I am being too pessimistic.”*

HCPs also expressed a desire to assess performance-based aspects of intelligence, such as processing speed. Examples of suggestions were to revise and adapt the ITVIK to an adult version or to develop a new test that could be administered in multiple stages to account for fatigue. Several HCPs emphasized the importance of linking intelligence assessment to practical functioning, advocating for a broader understanding of intelligence that goes beyond verbal and visual abilities.

##### HCP 3

> “*You focus less on what someone can do visually or verbally, which isolates specific functions. You cannot compensate. These subtests are designed to minimize the use of alternative capacities. But in practical settings, people often find other ways to manage… That too is a form of intelligence, in my opinion.”*

Two HCPs noted a lack of knowledge, either currently or in the past, regarding what is feasible in testing, and what viable alternatives exist to standard intelligence tests.

## Discussion

The aim of this mixed-method study was to get insight into the adaptations currently used during the assessment and interpretation of intelligence tests for adults with VI, based on international literature and input from Dutch clinical LVS practice.

Based on our literature review and interviews with HCPs, (subtests from) the WAIS-IV emerged as the most frequently administered intelligence test for adults with VI. Our literature review included tests that were developed or used within the past decade. In literature published prior to 2014, several intelligence tests for adults with VI were described. Rączy & Korczyk [2019] provide an overview of these instruments, including the Intelligence Test for Visually Impaired Children (ITVIC), especially developed to asses TIQ in children; Cognitive Test for the Blind (CTB), which measures cognitive function; the Paknikar, developed in India but has not found to be valid and reliable; or the B-101-DV which is only available in French (20). While these examples illustrate efforts to adapt intelligence testing for this population, they primarily target children or remain limited in scope and psychometric quality.

The main adaptations to the testing procedure, identified in this study, included administering only those subtests that are accessible to adults with VI, allowing extended time during testing, and using enlarged or haptic versions of test materials. Due to the heterogeneity of these adaptations, and concerns regarding the validity, reliability, and comparability of these adapted test, their results are expected to be compromised. Also, the literature does not describe any standardized protocols for selecting adaptations. Consequently, some HCPs develop their own small-scale organizational structures for this process.

HCPs emphasized that they took these adjustments closely into account during the interpretation of the results, and they documented these considerations in their reports. In the literature, only one study explicitly addressed the importance of accounting for such adaptations in the interpretation of results for adults with VI (34). They state that it is important to note the presence of a ceiling effect only for the blind participants in the present study. In addition, the British Psychological Society and Goodman et al. [2024] provide some practical advice for administering intelligence tests (39, 40). They advise test users to existentially describe adaptations in test usage, modifications to the environment, adaptations in interpretation, and observations and to incorporate these considerations into their explanations and feedback to the client.

Another mentioned adaptation to the testing procedure was the use of haptics; defined as tactilo-kinesthetic perception or the sense of active touch. A review of Mazella et al. [2014] found that haptics have been used in intelligence testing in two different ways (41). First, to assess nonverbal or practical intelligence, replacing vision in haptic analogues of mainstream tests (for example, tactile adaptations of Wechsler performance scales). And second, to assess, the quality of haptic functioning in specialized tests (for example, Tactual Profile, Haptic Test Battery) (42, 43). In the literature, only outdated tests, tests for children or tests that were not standardized for clinical use were found (41, 44, 45). A more recent example of haptics in intelligence testing is the Haptic Matrices Intelligence Assessment (HMIA) (46). Findings from a pilot study suggest that further refinement of the HMIA is required before it can serve as a valid measure of non-verbal intelligence for adults with VI, as its reliability was insufficient to meet the prerequisites for clinical application.

For the future, HCPs expressed a desire for a standardized protocol which could enhance reliability and reduce the influence of individual interpretation by ensuring consistency in test administration. The HCPs highlighted that current methods do not allow for reliable assessment of perceptual reasoning, working memory, and processing speed, making it difficult to accurately measure TIQ. HCPs report a need for measuring TIQ through specific intelligence tests and/or norm-groups tailored to adults with VI. Also in the literature review, no publications reported the use of adapted norm-groups. In line with our findings, previous research recommended that specific intelligence tests with specific norm-groups and dedicated manuals tailored to adults with VI should be developed (40, 47–49). This dedicated manual should outline procedures for describing functioning, handling test materials, and accounting for the impact of VI on test performance.

### Strengths and limitations

One notable strength of this mixed-methods study is the integration of a literature review and interviews conducted with HCPs working with adults with VI. This methodological triangulation enhances the robustness and credibility of the findings by drawing on insights from both research and clinical practice. Moreover, although the interviews were conducted at a national level, the challenges identified in measuring intelligence in adults with VI. It is likely generalizable and reflect broader, international issues as no intelligence tests in other languages are found in our literature review. The interviewees also had diverse professional experiences, which provided complementary perspectives: the long-term experience of senior HCPs offered valuable insights into test assessments, while the perspectives of newer HCPs contributed fresh and innovative ideas. Lastly, peer collaboration with two experts by experience was incorporated into the study design, further enhancing the relevance and practical applicability of the findings. Their involvement substantially strengthened the scientific rigor of the research through contributions to the test setup, the interview protocol, and their role as critical sparring partners for the researchers (50). The researchers expertise in intelligence testing in adults with VI added depth to the interviews, enabling more effective probing and elicitation of nuanced information from the HCPs.

The small-scale design of this study introduces certain methodological limitations. The literature review was restricted to publications from the past ten years, which may have led to the exclusion of older, potentially relevant studies. Nevertheless, this time frame ensured that the included publications reflecting current practice and reviews. However, when performing new studies, it is important to keep in mind earlier published intelligence tests which are currently nearly used or outdated. Additionally, due to time constraints, grey literature was not considered in the review, which could have led to missing suitable publications. The interviews were conducted within LVS organizations, however, the HCPs reported that they also receive inquiries from professionals outside the field of VI, including those working in mental health care. This highlights the broader relevance of their expertise, which in future research could provide critical insights into assessing adults with VI and inform best practices across multiple professional domains.

### Implications for future research and clinical practice

This study highlights the importance of developing tailored intelligence tests, appropriate norm groups, and/or standardized protocols to achieve accurate TIQ measurement and improve reliability for intelligence testing in people with VI. As long as there is no specific intelligence test to measure TIQ and/or norm-groups tailored to adults with VI, it is essential that all adaptations are described in detail, including the impact of the VI (40). This is of importance because HCPs without experience in the assessment of intelligence in adults with VI need to understand how to interpret the test scores similarly (51). Additionally, new research should focus on developing specific intelligence tests and norm-groups tailored to adults with VI. Hereby, developments of intelligence tests for other populations, such as for children with VI, can be studied.

Earlier research has shown that utilizing intelligence tests could put individuals at risk of misinterpretation of their cognitive abilities (52). For adults with VI this could be of importance when not all subtests can be reliable administered *i.e.* perceptual reasoning, working memory and processing speed. Therefore, when measuring the TIQ, HCPs should be aware of the potential for misinterpretation of test scores due to underestimation or overestimation resulting from knowledge sharing on this topic. Additionally, it will be important to continue research into what HCPs indicators are important to establish a TIQ-score in adults with VI.

Moreover, besides measuring a TIQ-score, the implications for individuals’ daily functioning could be taken into account. Currently, there is no systematic method available to accurately measure what adults with VI are actually capable of in their daily functioning. As mentioned by some HCPs, the ADAPT tool is an example of a test which could enable HCPs to observe individuals’ functioning in daily life. Further research is needed to study the assessment of and relationship between TIQ and practical intelligence. Practical intelligence could be particularly relevant because understanding a person’s abilities in daily practice helps tailor support and interventions to their real-world needs.

## Conclusion

Both literature and interviews with HCPs indicate that over the past decade, there has been no standardized, validated intelligence test available for adults with VI, highlighting a critical gap in psychodiagnostics testing for this group. Only a limited number of publications addressed intelligence testing in this population, and HCPs mentioned the absence of tailored tests and a variety of adaptations and interpretations. The WAIS-IV is the most frequently used test, in which often only verbal subtests are administered. In current practice, it seems impossible to obtain an accurate measure of TIQ for adults with VI. And therefore, a need is expressed for specific intelligence tests and norm-groups tailored to adults with VI. The importance of testing protocols for this population is emphasized, aiming to achieve a more uniform and standardized approach to assessment. The ability to accurately measure TIQ is essential to ensure that appropriate management strategies and support can be provided to adults with VI.

## Acknowledgement

The authors thank the interviewees from Robert Coppes Foundation, Bartiméus and Royal Dutch Visio, for their valuable contributions. Furthermore, they acknowledge the National Expert Network on VI and Psychiatry within Kennis over Zien, the Consortium for Expertise in Living with VI, and two experts by experience: Michel Gerritse and José Fijneman for their valuable feedback on the design of the study. Lastly, the authors thank S. Ghijsels (Brailleliga, Belgium) and L. Billiet (Amsterdam UMC) for carefully reading the manuscript and for comparing it with the current state of intelligence testing in Belgium, a country where a significant portion of the population speaks Dutch.

## Authorship and contributorship

M.O. and another registered psychologist formulated the research question from clinical practice. After funding was secured, C.L. developed the protocol. E.M., M.B., and M.O., with two experts by expertise, contributed to the design. C.L. conducted the literature search; C.L., E.M., and M.B. selected studies, with M.B. and M.O. resolving disagreements. Quality assessment and thematic analysis were carried out by C.L. and M.O. Interviews were conducted by M.O. The manuscript was drafted by C.L. and M.O. and revised by E.M., HA and M.B.

## Acknowledgement of AI use

Artificial Intelligence (AI), specifically ChatGPT and Copilot, were used to translate and enhance the English grammar and language of this manuscript. Moreover, it was used to translate one publication of the literature review from French to Dutch. Additionally, it was used to assist in finding relevant literature on the topic of the manuscript, while the authors reviewed all sources and remained fully responsible for the content.

## Data availability statement

Data are available upon reasonable request. Data consists of the article selection process and decisions made for inclusion in the review. Moreover, transcripts of interviews with healthcare professionals are available. Data is available under request of C. Lamper. No reuse without permission.

