## Supplementary file 1 for "Adaptations and interpretation of Intelligence Tests for Adults with Visual Impairment: A Literature Review and Interviews with Healthcare Professionals"

Supplementary file 1: Search Strategy

**1. Search Strategy for PubMed**

| **Concept** | **Keywords/Natural Language** | **MeSH Terms** |
| --- | --- | --- |
| **Intelligence tests** | "intelligence tests", "IQ tests", "cognitive assessments", "psychological testing", "Wechsler Adult Intelligence Scale (WAIS)", "Raven’s Progressive Matrices", "Stanford-Binet Intelligence Scales", "Woodcock-Johnson Tests of Cognitive Abilities", "Cattell Culture Fair Intelligence Test (CFIT)", "Kaufman Adult Intelligence Test (KAIT)", "Wonderlic Personnel Test", "General Aptitude Test Battery ( GATB)", "Shipley Institute of Living Scale (SILS)", "Leiter International Performance Scale", "Haptic Intelligence Scale for the Adult Blind (HISAB)", "Tactual Performance Test (TPT)", "Intelligence Test for Visually Impaired Persons (ITVIC)", "Braille-based Intelligence Tests", "Modified Bender-Gestalt Test", "Comprehensive Vocational Evaluation System (CVES)", "Adapted Torrance Tests of Creative Thinking", "Neale Analysis of Reading Ability" | "Intelligence Tests", "Cognition", "Neuropsychological Tests" |
| **AND** |  |  |
| **Use** | "use", "application", "administration", "implementation" | "Test Administration", "Psychological Test" |
| **OR** |  |  |
| **Interpretation** | "interpretation", "scoring", "evaluation", “measure” | "Test Interpretation", "Test Score Interpretation" |
| **AND** |  |  |
| **People with visual impairments** | "visual impairment", “Vision impairment” "blindness", "low vision", "vision loss", "visual disability" | "Visually Impaired Persons", "Blindness", "Vision Disorders" |

(Intelligence Tests[Mesh] OR Cognition[Mesh] OR Neuropsychological Tests[Mesh]) AND (intelligence tests OR IQ tests OR cognitive assessments OR psychological testing OR Wechsler Adult Intelligence Scale OR Raven’s Progressive Matrices OR Stanford-Binet Intelligence Scales OR Woodcock-Johnson Tests of Cognitive Abilities OR Cattell Culture Fair Intelligence Test OR Kaufman Adult Intelligence Test OR Wonderlic Personnel Test OR General Aptitude Test Battery OR Shipley Institute of Living Scale OR Leiter International Performance Scale OR Haptic Intelligence Scale for the Adult Blind OR Tactual Performance Test OR Intelligence Test for Visually Impaired Persons OR Braille-based Intelligence Tests OR Modified Bender-Gestalt Test OR Comprehensive Vocational Evaluation System OR Adapted Torrance Tests of Creative Thinking OR Neale Analysis of Reading Ability) AND (use OR application OR administration OR implementation OR scoring OR evaluation OR measure OR Test Administration OR Psychological Test OR Test Interpretation[Mesh] OR Test Score Interpretation[Mesh]) AND (visual impairment OR vision impairment OR blindness OR low vision OR vision loss OR visual disability OR "Visually Impaired Persons"[Mesh] OR "Blindness"[Mesh] OR "Vision Disorders"[Mesh])

Filter: 1/11/2014 - 1/11/2024

Imported References: 2580

**2. Search Strategy for Web of Science**

| **Concept** | **Keywords/Natural Language** | **Subject Headings (APA Thesaurus)** |
| --- | --- | --- |
| **Intelligence tests** | "intelligence tests", "IQ tests", "cognitive assessments", "psychological testing", "Wechsler Adult Intelligence Scale (WAIS)", "Raven’s Progressive Matrices", "Stanford-Binet Intelligence Scales", "Woodcock-Johnson Tests of Cognitive Abilities", "Cattell Culture Fair Intelligence Test (CFIT)", "Kaufman Adult Intelligence Test (KAIT)", "Wonderlic Personnel Test", "General Aptitude Test Battery (GATB)", "Shipley Institute of Living Scale (SILS)", "Leiter International Performance Scale", "Haptic Intelligence Scale for the Adult Blind (HISAB)", "Tactual Performance Test (TPT)", "Intelligence Test for Visually Impaired Persons (ITVIC)", "Braille-based Intelligence Tests", "Modified Bender-Gestalt Test", "Comprehensive Vocational Evaluation System (CVES)", "Adapted Torrance Tests of Creative Thinking", "Neale Analysis of Reading Ability" | "Intelligence Testing", "Cognitive Measurement", "Psychological Testing" |
| **AND** |  |  |
| **Use** | "use", "application", "administration", "implementation" | "Test Usage", "Assessment Administration" |
| **OR** |  |  |
| **Interpretation** | "interpretation", "scoring", "evaluation", “measure’’ | "Test Interpretation", "Score Interpretation" |
| **AND** |  |  |
| **People with visual impairments** | "visual impairment", "blindness", "low vision", "vision loss", "visual disability", “vision impairment” | "Visual Impairment", "Blindness", "Vision Loss" |

("Cognition" OR "Neuropsychological Tests" OR "intelligence tests" OR "IQ tests" OR "cognitive assessments" OR "psychological testing" OR "Wechsler Adult Intelligence Scale" OR "Raven's Progressive Matrices" OR "Stanford-Binet Intelligence Scales" OR "Woodcock-Johnson Tests of Cognitive Abilities" OR "Cattell Culture Fair Intelligence Test" OR "Kaufman Adult Intelligence Test" OR "Wonderlic Personnel Test" OR "General Aptitude Test Battery" OR "Shipley Institute of Living Scale" OR "Leiter International Performance Scale" OR "Haptic Intelligence Scale for the Adult Blind" OR "Tactual Performance Test" OR "Intelligence Test for Visually Impaired Persons" OR "Braille-based Intelligence Tests" OR "Modified Bender-Gestalt Test" OR "Comprehensive Vocational Evaluation System" OR "Adapted Torrance Tests of Creative Thinking" OR "Neale Analysis of Reading Ability") AND ("use" OR "application" OR "administration" OR "implementation" OR "scoring" OR "evaluation" OR "measure" OR "Test Administration" OR "Psychological Test" OR "Test Interpretation" OR "Test Score Interpretation") AND ("visual impairment" OR "vision impairment" OR "blindness" OR "low vision" OR "vision loss" OR "visual disability" OR "Visually Impaired Persons" OR "Blindness" OR "Vision Disorders")

Imported references: 50

**2. Search Strategy for PsycINFO**

| **Concept** | **Keywords/Natural Language** | **Subject Headings (APA Thesaurus)** |
| --- | --- | --- |
| **Intelligence tests** | "intelligence tests", "IQ tests", "cognitive assessments", "psychological testing", "Wechsler Adult Intelligence Scale (WAIS)", "Raven’s Progressive Matrices", "Stanford-Binet Intelligence Scales", "Woodcock-Johnson Tests of Cognitive Abilities", "Cattell Culture Fair Intelligence Test (CFIT)", "Kaufman Adult Intelligence Test (KAIT)", "Wonderlic Personnel Test", "General Aptitude Test Battery (GATB)", "Shipley Institute of Living Scale (SILS)", "Leiter International Performance Scale", "Haptic Intelligence Scale for the Adult Blind (HISAB)", "Tactual Performance Test (TPT)", "Intelligence Test for Visually Impaired Persons (ITVIC)", "Braille-based Intelligence Tests", "Modified Bender-Gestalt Test", "Comprehensive Vocational Evaluation System (CVES)", "Adapted Torrance Tests of Creative Thinking", "Neale Analysis of Reading Ability" | "Intelligence Testing", "Cognitive Measurement", "Psychological Testing" |
| **AND** |  |  |
| **Use** | "use", "application", "administration", "implementation" | "Test Usage", "Assessment Administration" |
| **OR** |  |  |
| **Interpretation** | "interpretation", "scoring", "evaluation", “measure’’ | "Test Interpretation", "Score Interpretation" |
| **AND** |  |  |
| **People with visual impairments** | "visual impairment", "blindness", "low vision", "vision loss", "visual disability", “vision impairment” | "Visual Impairment", "Blindness", "Vision Loss" |

(DE "Intelligence Tests" OR DE "Cognition" OR DE "Neuropsychological Tests" OR intelligence tests OR IQ tests OR cognitive assessments OR psychological testing OR Wechsler Adult Intelligence Scale OR Raven’s Progressive Matrices OR Stanford-Binet Intelligence Scales OR Woodcock-Johnson Tests of Cognitive Abilities OR Cattell Culture Fair Intelligence Test OR Kaufman Adult Intelligence Test OR Wonderlic Personnel Test OR General Aptitude Test Battery OR Shipley Institute of Living Scale OR Leiter International Performance Scale OR Haptic Intelligence Scale for the Adult Blind OR Tactual Performance Test OR Intelligence Test for Visually Impaired Persons OR Braille-based Intelligence Tests OR Modified Bender-Gestalt Test OR Comprehensive Vocational Evaluation System OR Adapted Torrance Tests of Creative Thinking OR Neale Analysis of Reading Ability)

AND (DE "Test Administration" OR DE "Psychological Tests" OR DE "Test Interpretation" OR DE "Score Interpretation" OR use OR application OR administration OR implementation OR scoring OR evaluation OR measure)

AND (DE "Visual Impairments" OR DE "Blindness" OR DE "Vision Disorders" OR visual impairment OR vision impairment OR blindness OR low vision OR vision loss OR visual disability OR visually impaired persons)

Publication Date: November 2014- October 2024

Results: 300

**3. Search Strategy for Google Scholar**

| **Concept** | **Keywords/Natural Language** | **Notes** |
| --- | --- | --- |
| **Intelligence tests** | "intelligence tests", "IQ tests", "cognitive assessments", "psychological testing", "Wechsler Adult Intelligence Scale (WAIS)", "Raven’s Progressive Matrices", "Stanford-Binet Intelligence Scales", "Woodcock-Johnson Tests of Cognitive Abilities", "Cattell Culture Fair Intelligence Test (CFIT)", "Kaufman Adult Intelligence Test (KAIT)", "Wonderlic Personnel Test", "General Aptitude Test Battery (GATB)", "Shipley Institute of Living Scale (SILS)", "Leiter International Performance Scale", "Haptic Intelligence Scale for the Adult Blind (HISAB)", "Tactual Performance Test (TPT)", "Intelligence Test for Visually Impaired Persons (ITVIC)", "Braille-based Intelligence Tests", "Modified Bender-Gestalt Test", "Comprehensive Vocational Evaluation System (CVES)", "Adapted Torrance Tests of Creative Thinking", "Neale Analysis of Reading Ability" | No MeSH or subject headings available; use natural language search terms and Boolean operators |
| **AND** |  |  |
| **Use** | "use", "application", "administration", "implementation" |  |
| **OR** |  |  |
| **Interpretation** | "interpretation", "scoring", "evaluation", “measure” |  |
| **AND** |  |  |
| **People with visual impairments** | "visual impairment", "blindness", "low vision", "vision loss", "visual disability", “vision impairment” |  |

("intelligence tests" OR "IQ tests" OR "cognitive assessments" OR "psychological testing" OR "Wechsler Adult Intelligence Scale" OR "Raven’s Progressive Matrices" OR "Stanford-Binet Intelligence Scales" OR "Woodcock-Johnson Tests of Cognitive Abilities" OR "Cattell Culture Fair Intelligence Test" OR "Kaufman Adult Intelligence Test" OR "Wonderlic Personnel Test" OR "General Aptitude Test Battery" OR "Shipley Institute of Living Scale" OR "Leiter International Performance Scale" OR "Haptic Intelligence Scale for the Adult Blind" OR "Tactual Performance Test" OR "Intelligence Test for Visually Impaired Persons" OR "Braille-based Intelligence Tests" OR "Modified Bender-Gestalt Test" OR "Comprehensive Vocational Evaluation System" OR "Adapted Torrance Tests of Creative Thinking" OR "Neale Analysis of Reading Ability")

AND ("use" OR "application" OR "administration" OR "implementation" OR "scoring" OR "evaluation" OR "measure" OR "test administration" OR "psychological test" OR "test interpretation" OR "test score interpretation")

AND ("visual impairment" OR "vision impairment" OR "blindness" OR "low vision" OR "vision loss" OR "visual disability" OR "visually impaired persons" OR "blindness" OR "vision disorders")

Aangepast bereik: 2014-2024

Resultaten: 66

**Appendix**

Exisiting intelligence tests for people with visual impairments.

**1. Wechsler Adult Intelligence Scale (WAIS) – Braille or Verbal Adaptation**

- **Overview**: The WAIS is commonly adapted for individuals with visual impairments by administering it verbally or using Braille versions of the test.
- **Adaptation**: For individuals who are blind or have low vision, tasks requiring visual input (such as block design or picture completion) are omitted or replaced with verbal reasoning tasks.
- **Use**: Suitable for individuals who are proficient in Braille or have a strong verbal ability.

**2. Verbal Intelligence Tests**

- **Overview**: Tests that focus primarily on verbal skills and do not require visual input are commonly used with visually impaired individuals.
- **Examples**:
  - **Wechsler Verbal Scales**: Only the verbal subtests of the WAIS, such as vocabulary, similarities, arithmetic, and comprehension, are used.
  - **Peabody Picture Vocabulary Test (PPVT)**: Administered verbally by describing the pictures or adjusting the format to avoid visual dependence.
- **Use**: Ideal for individuals with good auditory or verbal skills.

**3. Haptic Intelligence Tests (Tactual Performance Tests)**

- **Overview**: These tests are designed to assess intelligence through tactile methods, where individuals use touch rather than sight to complete tasks.
- **Examples**:
  - **The Haptic Intelligence Scale for the Adult Blind (HISAB)**: A test specifically developed for individuals who are blind, which includes tasks that can be completed through touch, such as identifying shapes or objects.
  - **Tactual Performance Test (TPT)**: Part of the Halstead-Reitan Neuropsychological Battery, this test assesses tactile perception and memory through tactile and motor skills tasks.
- **Use**: Suitable for individuals who rely on their sense of touch due to vision loss.

**4. Raven’s Progressive Matrices – Tactile Version**

- **Overview**: Raven’s Progressive Matrices is a nonverbal test of abstract reasoning that has been adapted into a tactile version for individuals with visual impairments.
- **Adaptation**: The visual patterns are converted into raised shapes or objects that can be felt, and participants are asked to identify the missing piece based on tactile input.
- **Use**: Ideal for individuals with strong tactile discrimination and problem-solving abilities.

**5. Intelligence Test for Visually Impaired Persons (ITVIC)**

- **Overview**: This is a specially designed intelligence test for visually impaired individuals.
- **Structure**: The test uses both verbal and tactile tasks to measure cognitive abilities such as verbal comprehension, working memory, and reasoning.
- **Use**: Specifically developed to accommodate adults with visual impairments, it avoids tasks that require sight.

**6. Braille-based Intelligence Tests**

- **Overview**: For individuals who are proficient in Braille, some intelligence tests have been adapted into Braille, allowing them to complete tasks through reading tactile symbols.
- **Examples**:
  - **Stanford Achievement Test – Braille Edition**: Used more for academic performance but can assess intelligence-related skills such as reasoning and comprehension.
  - **WAIS-Braille**: An adapted version of the WAIS for Braille readers.

**7. Modified Bender-Gestalt Test (for Tactile Perception)**

- **Overview**: Originally a visual-motor test, this test has been adapted for tactile perception to assess individuals with visual impairments.
- **Adaptation**: The visual patterns of the original test are replaced with raised shapes that the individual traces or feels to replicate using touch.
- **Use**: Assesses cognitive and perceptual motor skills using tactile rather than visual input.

**8. Comprehensive Vocational Evaluation System (CVES)**

- **Overview**: This is a nonverbal, hands-on assessment used to evaluate vocational and cognitive abilities in individuals with visual impairments.
- **Structure**: The test consists of a variety of hands-on tasks that assess reasoning, memory, and problem-solving through tactile methods.
- **Use**: Often used in vocational rehabilitation settings to assess cognitive strengths and job-related skills for individuals with visual impairments.

**9. Adapted Torrance Tests of Creative Thinking (Torrance Tactile Tests)**

- **Overview**: These tests assess creativity and problem-solving and have been adapted for tactile perception.
- **Structure**: Tasks focus on creative thinking and reasoning without relying on visual cues, using raised objects and patterns.
- **Use**: Useful for individuals with visual impairments to assess creative problem-solving.

**10. Neale Analysis of Reading Ability – Modified Version for the Blind**

- **Overview**: Primarily a reading ability test, but also used to assess comprehension and cognitive skills.
- **Adaptation**: Modified to be administered in Braille or through audio for individuals with visual impairments.
- **Use**: Measures intellectual ability related to reading and comprehension.
