## Supplementary file 2 for "Adaptations and interpretation of Intelligence Tests for Adults with Visual Impairment: A Literature Review and Interviews with Healthcare Professionals"

Supplementary file 2: Topic Guide
*Interview with Behavioral Scientists on Intelligence Assessment for People with Visual Impairment utilized between* *June 16, 2025, and October 6, 2025.*

**1. Introduction and Background (5 min)**

- Brief introduction of the researcher and the purpose of the interview
- Introduction of the interviewee:
  - Background
  - Education (pedagogical scientist, applied psychologist, basic psychologist, registered healthcare psychologist, or registered clinical psychologist)
  - Expertise in (neuro) psychological assessment
  - Experience with people with visual impairment (VI)

***Aks for oral informed consent + recording – always the option to stop***

**2. Use of Intelligence Assessment in Adults with Visual Impairment (15 min)**

- What is your experience with administering intelligence assessments in individuals with VI?
- And what differences do you observe compared to administration in sighted individuals?
- Which assessments are utilized?
- How frequently?
- How is reliability, validity, and quality taken into account? Ask for knowledge of the COTAN.
  *(COTAN: The mission of COTAN is to promote the quality of tests and test use in the Netherlands by informing test users, test developers, and test publishers about the availability, content, and quality of various instruments. It does so by evaluating the quality of a wide range of tests, examinations, and questionnaires, and by establishing standards for the use of psychological instruments, such as the General Standard for Test Use by NIP.)*
- Which criteria do you apply when selecting a specific intelligence assessment?
- Are certain assessments considered more or less suitable? Why?
- Which guidelines or protocols exist for administering intelligence assessments to individuals with VI?
- Has your organization developed its own guidelines or protocols for administering intelligence assessments?
- Does the administration (of the subtests administered) take as long as with sighted individuals?

**3. Adaptations and Assessment Methods (10 min)**

- Which adaptations do you make, or would you make, in the administration of intelligence assessments for individuals with VI?
  - Which components of the IQ assessment do you include?
  - Which do you exclude?
  - What adaptations are applied?
- What are the advantages and limitations of these adaptations?
- How do these adaptations influence test performance and interpretation? In what way do you report and communicate this?
- What is your opinion on providing additional time?
- Do you differentiate between client groups?
  - Percentage of vision?
  - Acquired vs. congenital?

*In the literature, we see that intelligence scores are often determined based on verbal components, excluding visual components. We also observe that subsets of intelligence assessments are frequently administered to map cognitive abilities, without calculating a full IQ score.*

- What is your perspective on this?
- Do you consider this approach sufficient to provide an overview of a client’s capacities? If yes, why? If not, why not—and what would be needed to obtain such an overview?
- What are your experiences with tactile or verbal alternatives (such as Braille versions or verbal subtests) in intelligence assessment?

**4. Interpretation and Norming (10 min)**

- How do you interpret assessment results for individuals with VI?
- How reliable and valid are these results for this population?
- How do you and your colleagues norm the assessment outcomes?
- Do you use specific methods tailored VI, or norming for tactile or verbal subtests?
- How do you report the adaptations made compared to standard administration?
- Are there adapted norm groups, or would such groups be necessary? Why is this important, or not?
- How do you communicate the assessment results to clients and their network?
- Were there cases where a test score led to incorrect assumptions about someone’s abilities or potential?
- How can interpretation of assessment results be improved to accurately reflect the skills of individuals with VI?

**5. Need for Guidelines and Further Development (5 min)**

Returning to the earlier question on protocols (presence or absence), follow-up questions:

- What knowledge do you currently lack regarding intelligence assessment in individuals with VI?
- If standardization of intelligence assessments were introduced within the VI sector, what should this look like for you?
- What elements should definitely be included or excluded?
- How would standardization/protocols/guidelines for intelligence assessment influence your administration process?

**6. Significance of Assessment Results for the Client (10 min)**

- How have clients for whom you administered an intelligence assessment experienced the process?
  - Cognitive demanding? Fatiguing?
- How do clients perceive the outcome of the intelligence assessment?
  - Unjustly underestimated or overestimated?
  - Impact on self-image? Feelings of self-worth?

**7. Closing and Additional Remarks (5 min)**

- Are there any important aspects that have not been discussed but are relevant to this topic?
- Thank the interviewee and confirm consent for the possibility of follow-up contact.
