## Supplementary file 3 for "Adaptations and interpretation of Intelligence Tests for Adults with Visual Impairment: A Literature Review and Interviews with Healthcare Professionals"

### Supplementary file 3: Overview Intelligence Tests Mentioned in Publications and/or Interviews

| **Name test** | **Purpose** | **Target group** | **Subtests** | **Subtests applied in publications** | **Test mentioned in x publications** | **Test mentioned in x interviews** |
| --- | --- | --- | --- | --- | --- | --- |
| Wechsler Adult Intelligence  Scale-fourth edition *(WAIS-IV)* | Assessing adult’s and older adolescent’s cognitive abilities in clinical, educational, and research settings. | Adults from 16:0 to 90:11 years | 10 core subtests and 5 optional subtests, grouped into four index scores.  Verbal Comprehension Index Core: Similarities, Vocabulary, Information Optional: Comprehension  Perceptual Reasoning Index Core: Block design, Matrix reasoning, Visual puzzles  Optional: Picture completion, Figure weights  Working Memory Index Core: Digit span, Arithmetic Optional: Letter-number sequencing  Processing Speed Index Core: Symbol search, Coding Optional: Cancellation | Digit Span (N=4)  Letter-Number Sequencing (N=1)  Tests from Verbal Comprehension Scale  (N=1)  Not specified (N=2) | N = 6 | N = 9 |
| Wechsler Intelligence Scale  for Children -fifth edition *(WISC-V)* | Assessing children’s cognitive abilities in clinical, educational, and research settings. | Children from 6:0 to 16:11 years | 10 core subtests and 5 optional subtests, grouped into five index scores  Verbal Comprehension Index  Core: Similarities, Vocabulary  Optional: Information, Comprehension  Visual Spatial Index  Core: Block Design, Visual Puzzles  Fluid Reasoning Index  Core: Matrix Reasoning,  Figure Weights  Optional:  Picture Concepts  Working Memory  Index  Core: Digit Span  Optional: Letter-Number Sequencing  Processing Speed Index  Core: Coding, Symbol Search  Optional: Cancellation | Not specified | N = 2 | N=4 |
| Intelligence test for visually impaired children (ITVIK)* | Dutch test to assess cognitive abilities in children with visual impairments | Children from 6:0 till 15:0 years | 13 verbal and tactile (haptic) subtests based on Thurstone’s model of intelligence | N=0 | N=0 | N=5 |
| Intelligence and Development Scales – Second Edition  *(IDS-2)* | Brief psychological assessment used to evaluate intelligence and developmental functioning in children and adolescents | Children and adults from 5:0 till 20:0 years | Intelligence domains  Verbal reasoning, Non-verbal / fluid reasoning, Visual–spatial abilities, Working memory, Processing speed  Developmental domains Executive functions, Psychomotor skills, Academic skills (reading, spelling, mathematics), Social-emotional competencies (via supplementary scales) | N=0 | N=0 | N=1 |
| Paknikar | Intelligence in people with VI | Children from 8:0 to 18.0 years, adaptable for older individuals | Comprehension, Memory, and Reasoning using tactile and kinaesthetic performance tasks. | N=0 | N=0 | N=1 |
| Wechsler Abbreviated Scale of Intelligence-second edition  *(WASI-II)* | Brief, reliable measure of intelligence for clinical, educational, and research use | Children and adults from 6:0 to 90:11 years | Verbal Comprehension Index Similarities, Vocabulary  Perceptual Reasoning Index Block design, Matrix reasoning  FSIQ-4 and FISQ-2 scores  Estimate of general cognitive ability based on the four or two (Vocabulary, Matrix reasoning) tests | Not specified (N=1) | N=1 | N=0 |
| Dutch Adult Reading Test *(DART)* | Estimation of premorbid IQ | 25:0 to 80:0 years | Verbal Components  Single-word reading test | Single-task test N=1 | N=2 | N=0 |
| Stanford-Ohwaki-Kohs Tactile Block Design Intelligence Test | Nonverbal/performance intelligence (tactile spatial reasoning), comparable to the Wechsler Block Design subtest, but tactile. | Blind adolescents and adults from 14:00 years and older | Nonverbal Intelligence  Tactile problem-solving | Single-task test N=1 | N=1 | N=0 |
| Test of Nonverbal Intelligence-fourth edition *(TONI-4)* | General intelligence especially for those with speech, language, hearing or motor impairments, or in case of diverse cultural and linguistic backgrounds | Children and adults from 6:00 to 89:11 years | Nonverbal Intelligence and Abstract reasoning Unidimensional test composed of 60 abstract-figural items | Not specified N=1 | N=1 | N=0 |
| Otis Classification Test-Revised *(OTIS‑R)* | General intelligence and academic achievement | Children from $\pm$ 9:00 to 15:00 years | General Intelligence Verbal tasks: Analogies, Sentence completion, Vocabulary and definitions, Logic reasoning using language, Reading comprehension questions  Nonverbal reasoning tasks: Pattern recognition, Series completion, Classification of shapes or symbols, Spatial reasoning, Matching based on visual attributes  Academic Achievement School subjects: Reading, Spelling, Languare, Arithmetic and fundamentals, Social studies, Health, General information | Not specified N=1 | N=1 | N=0 |
| Similarities-R | Verbal abstraction and categorisation | Not specified | Similarities | Not specified N=1 | N=1 | N=0 |
| Arithmetic-R | Mathematical reasoning, concentration and concept manipulation | Not specified | Arithmetic | Not specified N=1 | N=1 | N=0 |
| Raven’s Progressive Matrices-R | Fluid and abstract reasoning | Up to 4:00 years till 70+ years | Simple pattern completion, Analogies, Abstract relationships, Complex logical progressions and Multiple rules combined | Not specified N=1 | N=1 | N=0 |
| Haptic Intelligence Scale for the Blind  *(HISB)* | Non-verbal intelligence | 16:0 till 64:00 years | Digit Symbol, Block Design, Object Assembly, Object Completion, Pattern Board, Bead Arithmetic | Not specified N=1 | N=1 | N=0 |
| B-101 DV | Assessment of general intelligence in children with VI, adapted to their visual limitations | Children from 6:0 till 16 years | Verbal reasoning Vocabulary, Comprehension, Verbal problem-solving tasks  Nonverbal reasoning  **Tactile or haptic tasks** – Arranging shapes, patterns, or objects to assess nonverbal reasoning.  **Memory tasks** – recalling sequences or patterns presented through touch or auditory means.  **Analogy and classification tasks** – using tactile materials or verbal descriptions. |  | N=1 | N=0 |

* The Intelligence Test for Visually Impaired Children (ITVIK) is currently being under revision.
