## Supplementary file 4 for "Adaptations and interpretation of Intelligence Tests for Adults with Visual Impairment: A Literature Review and Interviews with Healthcare Professionals"

Supplementary file 4: Critical appraisal

Supplementary File 3: Critical appraisal of each included study using the JBI Critical Appraisal Checklists

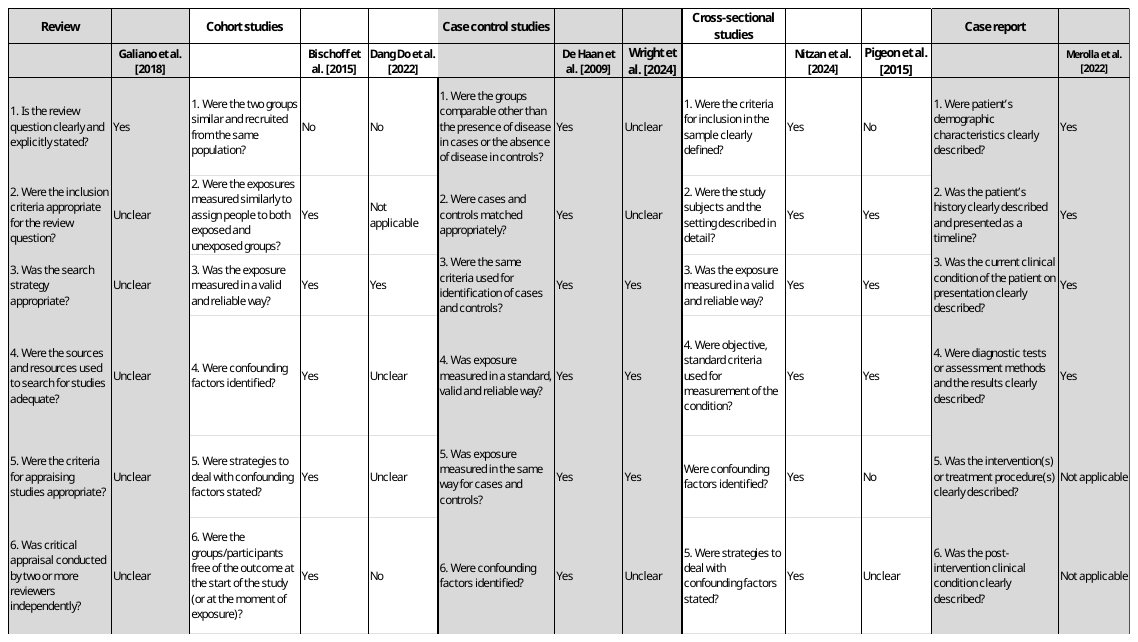

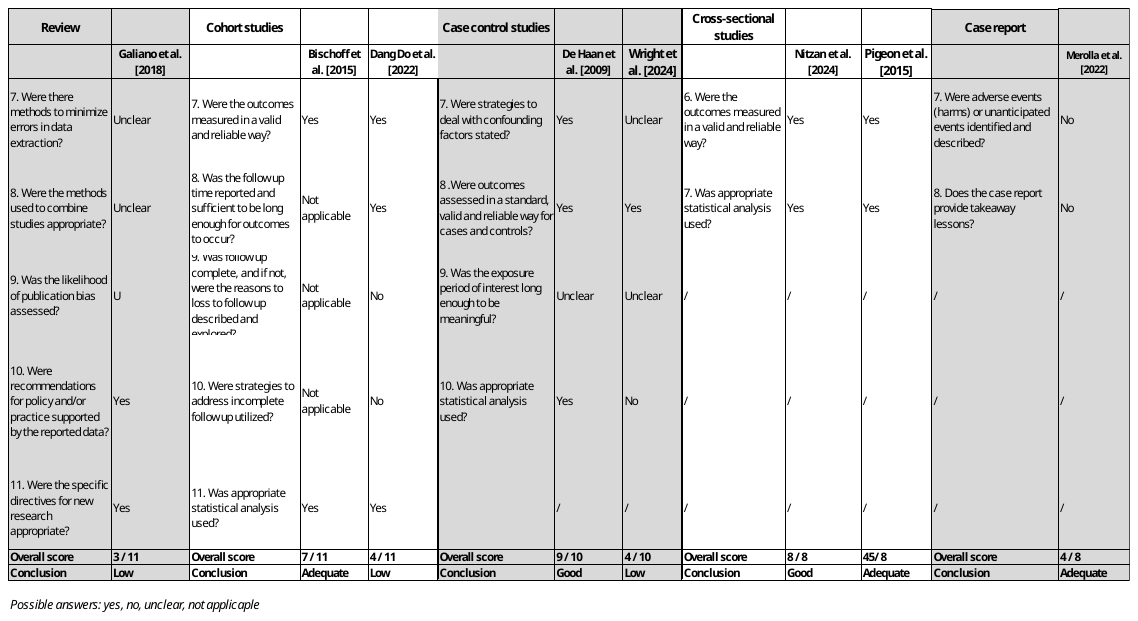
